# Puerto Rico Health System Resilience After Hurricane Maria: Implications for Disaster Preparedness in the COVID-19 Era

**DOI:** 10.1101/2020.09.20.20198531

**Authors:** Christopher C. Rios, Emilia J Ling, Ralph Rivera-Gutierrez, Juan Gonzalez Sanchez, Janine Bruce, Michele Barry, Vinicio de Jesus Perez

## Abstract

**Background:** Every year, Puerto Rico faces a hurricane season fraught with potentially catastrophic structural, emotional and health consequences. In 2017, Puerto Rico was hit by Hurricane Maria, the largest natural disaster to ever affect the island. Several studies have estimated the excess morbidity and mortality following Hurricane Maria in Puerto Rico, yet no study has comprehensively examined the underlying health system weaknesses contributing to the deleterious health outcomes.

**Methods:** A qualitative case study was conducted to assess the ability of the UPR health system to provide patient care in response to Hurricane Maria. An established five key resilience framework and inductive analysis was used to identify factors that affected health system resilience. Thirteen Emergency Medicine Physicians, Family Medicine Physicians, and Hospital Administrators in a University of Puerto Rico (UPR) Community Hospital were interviewed as part of our study.

**Results:** Of the five key resiliency components, three domains were notably weak with respect to UPR’s resiliency. Prior to the Hurricane, key personnel at the UPR hospital were *unaware* of the limited capacity of back-up generators at hospitals and were ill-prepared to transfer ICU patients to appropriate hospitals. Post Hurricane, the hospital faced *self-regulation* challenges when triaging the provision of Hurricane-related emergency services with delivering core health services, in particular for patients with chronic conditions. Finally, during and after the Hurricane, *integration* of patient care coordination between the UPR hospital ambulances, neighboring hospitals, and national and state government was suboptimal. The two remaining resiliency factors, addressing *diverse* needs and system *adaptiveness* in a time of crisis, were seen as strengths.

**Conclusions:** Hurricane Maria exposed weaknesses in the Puerto Rican health system, notably the lack of awareness about the limited capacity of backup generators, poor patient care coordination, and interruption of medical care for patients with chronic conditions. As in other countries, the current COVID epidemic is taxing the capacity of the Puerto Rico health system, which could increase the likelihood of another health system collapse should another hurricane hit the island. Therefore, a resilience framework is a useful tool to help health systems identify areas of improvement in preparation for possible natural disasters.

## Introduction

In September 2017, Hurricanes Irma and Maria devastated Puerto Rico and caused severe damage to critical infrastructure and essential health systems [1; 2]. This led to an ongoing, massive shortage of electricity and clean water supplies, which had substantial impact on the health and well-being of the Island’s population. Although disruption in healthcare services is expected during any natural disaster, health systems must be able to minimize these disruptions and respond to these stressors in a time of need [3; 4; 5].

Recently, there has been a growing body of literature characterizing the importance of resilience in health systems. Now, with the spread of COVID-19 challenging the integrity of health systems around the globe, this concept has once again been brought to the forefront. Resilience, defined as the capability of a health system to prepare, respond and reorganize under conditions of stress, is posited to protect the population from excess morbidity and mortality [3; 5; 6; 7; 8; 9]. In this setting, excess mortality and morbidity is defined as an elevation in the number of deaths and/or prevalence of disease in the months to years following the stressor as compared to the expected number of deaths and/or prevalence of disease if the stressor had not occurred. The most comprehensive assessment of Hurricane Maria found that from September 20, 2017 to December 31, 2017, there was an estimated 4,645 excess deaths largely due to delayed or inadequate access to healthcare services[2]. Moreover, various studies have shown that people with chronic medical conditions, and persons in the lowest socioeconomic category were disproportionately affected by Hurricane Maria [1; 2; 10; 11].

Although there has been extensive literature focusing on the definition of resilience in health systems, there have been few studies which apply the resilience framework to health systems [8; 12]. In this study, a qualitative approach is used to bring the resilience framework from the theoretical to the practical. Specifically, a resilience framework is used to identify key factors that may have contributed to excess morbidity and mortality in Puerto Rico following Hurricane Maria.

## Methods

### Setting, context, and rationale

We conducted a case study using qualitative methods, as this approach allowed us to better understand the ‘why’ and ‘how’ of excess mortality and morbidity following Hurricane Maria [13]. The UPR Community Hospital was purposefully selected as the ‘critical case’ given its central location in San Juan (the capital of Puerto Rico), because it was one of the few hospitals that remained open throughout Hurricane Maria and the critical weeks following the natural disaster, and it has disaster-preparedness systems similar to other hospitals. Due to these characteristics, this ‘critical case’ case study about this UPR Community Hospital’s resilience may help to identify successful strategies that other hospitals/healthcare systems may consider when implementing policies to address resilience factors [13; 14].

### Participants and approach

To assess hospital and health system resilience, semi-structured in-depth interviews were conducted with key stakeholders at the selected UPR Community Hospital (**Table 1**). We used key informant sampling of physicians who were on service for at least three days in the week before or after Hurricane Maria. To capture a breadth of understanding and experiences, we identified Emergency Medicine (EM) physicians, Family Medicine (FM) physicians, and Administrators involved in delivering or coordinating patient care before, during or after Hurricane Maria. The interviews generally lasted between 30 and 70 minutes. Reflexivity was maintained by researchers throughout the interview and analysis process by recording, discussing, and challenging established assumptions throughout the data collection and analysis.

**Table 1:** Demographics of Respondents.

|  | **Subjects**<br>**(n = 13)** | **Percentage** |
| --- | --- | --- |
| **Male/Female** | 5/8 | 38.5/61.5 |
| **Years of Experience:** |  |  |
| 0-1 | 3 | 23.1 |
| 1-2 | 6 | 46.2 |
| 3+ | 4 | 30.8 |
| **Hospital Affiliation:** |  |  |
| Emergency Medicine (EM) | 4 | 30.8 |
| Family Medicine (FM) | 4 | 30.8 |
| Administration | 5 | 38.5 |
| **Time Spent at Hospital:** |  |  |
| 1-3 Days pre-Hurricane | 9 | 75.0 |
| During Hurricane | 10 | 83.3 |
| 1-3 Days post-Hurricane | 11 | 91.7 |
| Only After | 2 | 16.7 |

An interview guide was created by the first and second author adapted from a similar study conducted in Ebola by the second author [5]. The interview guide was designed to prompt discussion about the strengths and weaknesses the participants observed after and before the Hurricane through the lens of the five resilience domains. The questions were revised and adapted to the cultural context, then reviewed by two qualitative research experts for integrity. Interviews were conducted between June 15, 2018 and August 5, 2018. All interviews were conducted in-person with identified stakeholders.

Confidentiality and anonymity were discussed with participants during the consent process. Interviews were audiotaped for transcription and purposes conducted in both Spanish and English depending on the preference of the participant. All study findings, including quotations, have been appropriately anonymized, and data collected through this study was safely stored in a password-protected laptop. The Stanford Institutional Review Board approved all research procedures as an exempt study.

### Data Analysis

Using the resilience framework as the initial codes, we conducted deductive thematic analysis to identify major themes that impacted hospital and health system resilience during the response to Hurricane Maria [5]. For our starting domains, we used the Kruk framework of health system and hospital resilience. Under this framework, health system or hospital resilience is broken down into its five foundational components: (1) Aware, (2) Diverse, (3) Self-Regulating, (4) Integrated, and (5) Adaptive [5]. **Table 2** summarizes the foundational components and the characteristics of resilient hospitals and health systems.

**Table 2:** Health System Resilience Domains.

| **Domain** | **Definition** |
| --- | --- |
| **Aware** | Obtains a comprehensive, up-to-the-minute understanding of the specific health needs of the population as well as the human, physical, technical assets necessary to meet those health needs |
| **Diverse** | Meets a substantial breadth of the population's health needs |
| **Self-Regulating** | Continues to meet the basic health needs of the population when the hospital or system is under significant stress |
| **Integrated** | Effectively and efficiently brings together diverse groups of actors to coordinate patient care and deliver health services |
| **Adaptive** | Transforms when under stress to meet the specific health needs of the population |

After using the Kruk resilience framework as initial codes, we conducted iterative inductive analysis to identify additional resilience themes that arose from the data. Inter-rater reliability was achieved via Dedoose training to ensure credibility of the results [15]. Specific resilience themes were identified through data familiarization by reading and rereading coded excerpts. Through this method, various factors of health system resilience were identified that participants expressed likely contributed to excess morbidity and mortality. Finally, this study fulfills standards for reporting qualitative research [16; 17].

### Research reporting checklist

We used the SRQR reporting guideline (O’Brien BC, Harris IB, Beckman TJ, Reed DA, Cook DA. Standards for reporting qualitative research: a synthesis of recommendations. Acad Med. 2014;89(9):1245-1251.)

## Results

In the following section, we report on the five resilience framework categories and the inductively derived themes/sub-themes that emerged in the context of Hurricane Maria. These are summarized in **Table 3**.

**Table 3:**
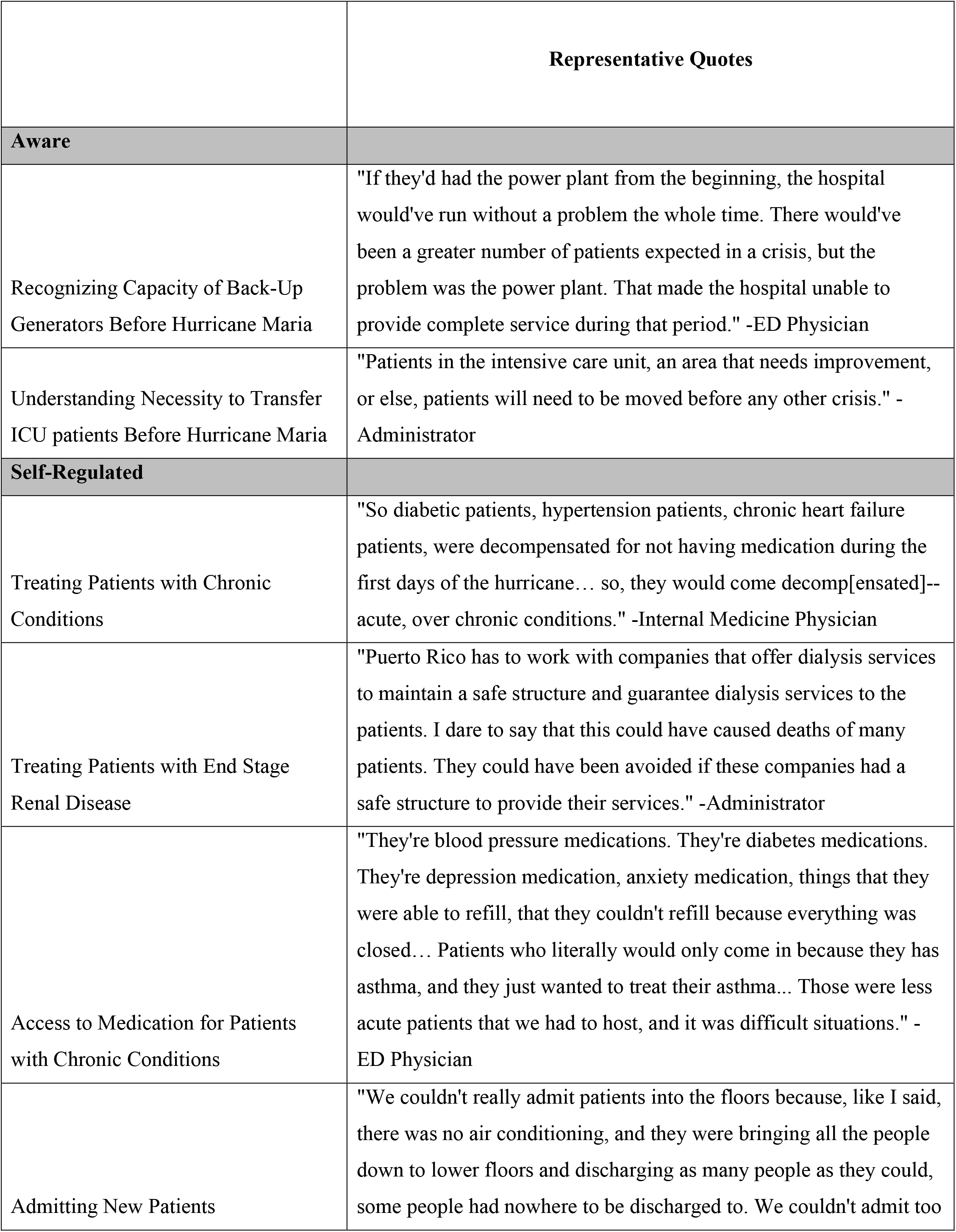
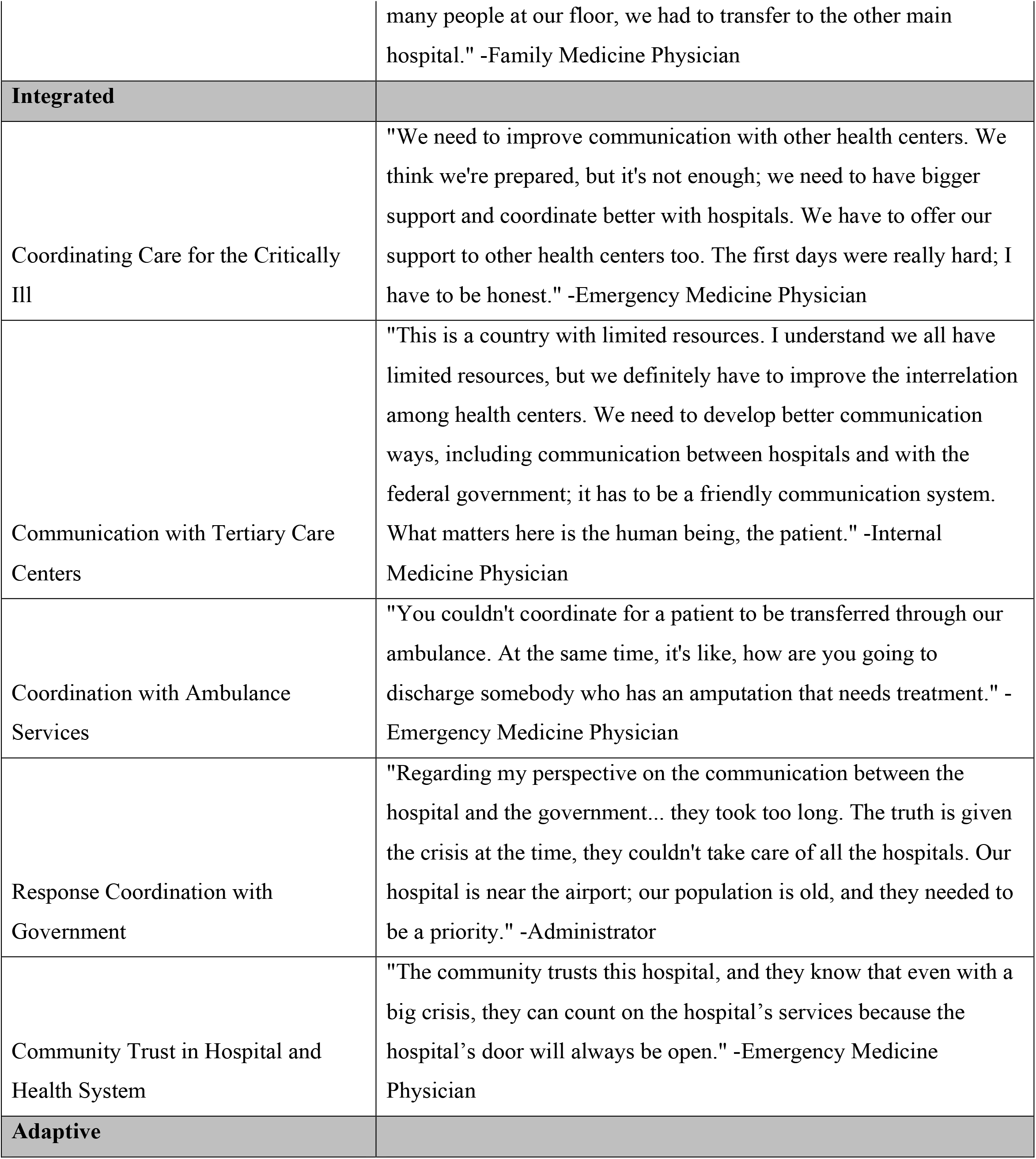
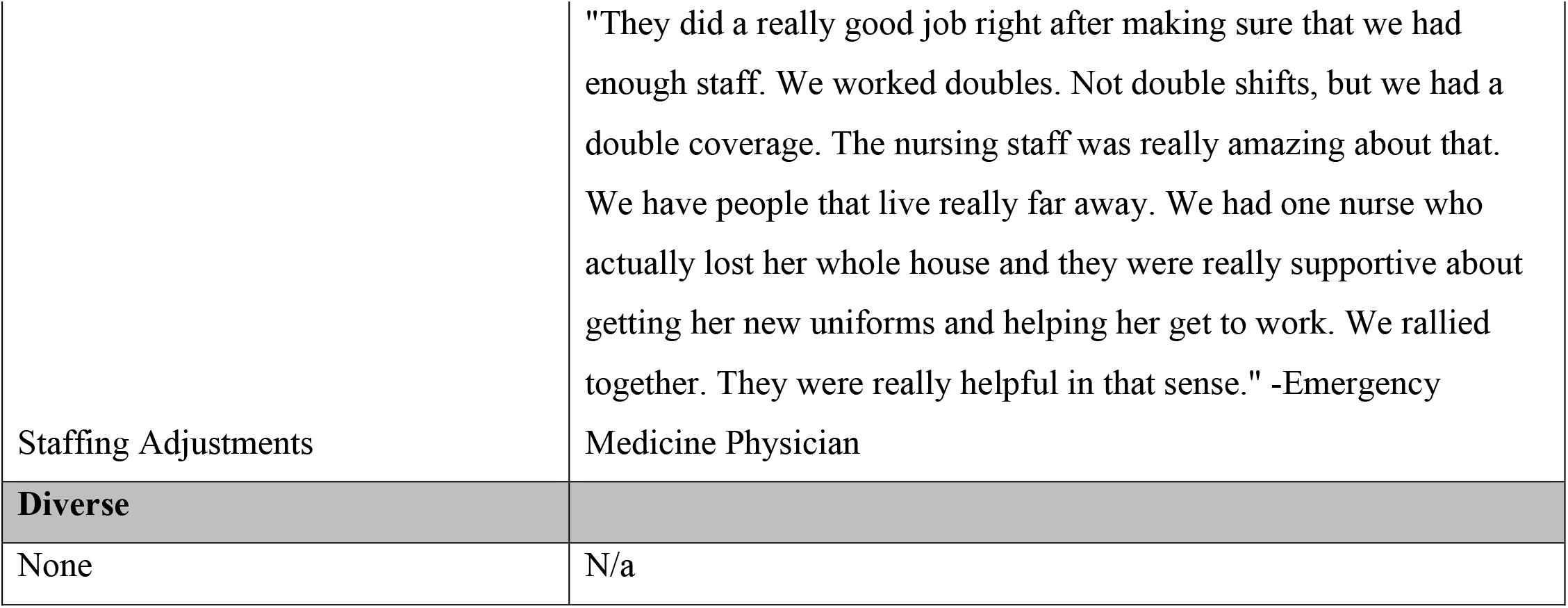
Hospital/Health System Themes and Sub-themes with Representative Quotes.

### Aware

#### Recognizing Limited Capacity of Emergency Back-Up Generators before Hurricane Maria

Physicians and administrators described the lack of back-up generators affecting patient care in a variety of ways, and many described it as the single greatest factor contributing to interruption of patient care. One major problem was the lack of temperature regulation for very ill patients. As one EM physician describes, “I think one issue was the heat. The patients that we weren’t able to bring down to floors that were cooler, their family brought them a fan. So, it was really, really hot and, in part, that affected mortality. I feel like the heat was part of it.” Another FM physician described the difficulty of treating patients in labor and delivery without power, “It was very hot, and humid—but we tried to maintain the best possible habitat for the pregnant ladies. But there was a point where the labor room—wasn’t able to take any more patients, that was an order of the medical director because— it was not okay to work in those conditions.” Patients had to be moved from floors without electricity because of the heat, which resulted in significant overcrowding of the emergency room and lobby. To many of the respondents, the power supply was the ultimate limiting factor when it came to providing adequate care, “The power supply was our limitation. We needed a power generator with enough capacity to supply the entire hospital. There was no oxygen, and patients were very hot. If the patient had an ulcer, the ulcer got worse.”

#### Understanding Necessity to Transfer ICU Patients before Hurricane Maria

Given the constraints on power supply, this UPR Community Hospital was unable to provide adequate care for patients in the ICU and many of these patients had to be chaotically transferred to other hospitals in the days following the hurricane. One administrator noted, “We need to identify the hospital’s vulnerable areas to protect the patient in those areas or evacuate them prior to the disaster.” Another administrator noted the difficulty transferring these patients following the Hurricane, “It was difficult to transfer patients to be admitted to other health centers. It wasn’t easy to write a memo or call and say, ‘I have 20 patients that need to be moved.’ In the end we made it, but we need to improve this part.”

### Self-Regulated

#### Treating Patients with Chronic Conditions

The two populations that were most adversely affected were patients with end stage renal disease (ESRD) and patients that needed access to medications for chronic conditions. These findings corroborate other studies that suggest elderly populations with chronic conditions were disproportionately affected by Hurricane Maria [2; 18; 19].

#### Limited Treatment for Patients with End Stage Renal Disease (ESRD)

Patients with ESRD rely on regular hemodialysis to avoid life-threatening metabolic derangements such as hyperkalemia or uremia[20]. Given the prevalence of diabetes in Puerto Rico it is no surprise that there is a significant population with ESRD reliant on regular hemodialysis [11]. In the weeks following Hurricane Maria, these patients experienced extreme difficulty accessing treatment. As one EM physician stated, “As far as chronic patients, kidney failures were the most frequent visitors to the hospital after the event because they hadn’t been able to get their dialysis during those days.” Respondents noted that in the days and weeks following Hurricane Maria, decompensated patients with ESRD presented to the emergency department for emergent dialysis, “We had to deal with patients who had to be dialyzed urgently, so I think that having an additional dialysis unit for emergencies to help those patients could be an improvement.” In the aforementioned cases, patients were able to ultimately seek medical attention, but it is likely that many other patients with ESRD, especially those living in remote areas, were not able to access care.

#### Limited Access to Medications for Patients with Chronic Conditions

Patients with other chronic conditions like COPD, asthma, heart failure and others who also rely on consistent medical management to ensure that they do not decompensate into a life-threatening state, were disproportionally affected by Hurricane Maria. One administrator noted, “Diabetic patients, hypertension patients, chronic heart failure patients, were decompensated for not having access to mediation during the first few days of the hurricane. So they would come in decompensated—acute over chronic conditions.” Another respondent noted the difficulty addressing this situation, “There are no health centers for patients who need this type of long-time treatments. We didn’t have the resources because there were not that many beds.” Although many of the patients were ultimately able to receive respiratory treatment or access to their medications, many patients in remote regions of Puerto Rico were not able to travel to an emergency department to receive therapy. Moreover, even for patients that did manage to ultimately access their medications, acute exacerbation of chronic conditions often lead to increasing morbidity and more rapid disease progression.

#### Reduced Admission of New Patients

Respondents noted that, shortly after Hurricane Maria, the hospital was no longer able to admit new patients. As one EM physician noted, “We couldn’t admit patients so the challenge was: should we close our doors to these patients because we can’t offer them the services that they most deserve?” This created a chaotic situation where acutely ill patients had to be managed exclusively in the emergency department or sent away. Another EM physician noted, “If a patient with an infected wound, who has to be admitted to the hospital, came in, that patient wouldn’t be admitted.” Ultimately, these patients would be transferred to another hospital, but the inability to admit represented a significant delay in medical care.

### Integrated

#### Coordinating Care for Critically Ill

One of the most significant challenges encountered by the UPR Community Hospital following Hurricane Maria was coordinating care for the critically ill. As one EM physician noted, “The coordination, on the first day, the first days, the first weeks, it was nearly impossible because there was no communication.”

#### Broken Communication with Tertiary Care Centers

Immediately following Hurricane Maria, many patients had to be transferred from the UPR Community Hospital to Tertiary Care Centers. During regular practice, the UPR Community Hospital calls ahead to ensure that space is available and inform the hospital service that the patient is being transferred. This process allows for efficient and safe transfer of patients from one care center to another. In the days after Hurricane Maria, due to the interruption of all communication channels, patients were transferred blindly. One EM physician noted, “When we would receive trauma patients, we had to send them to the level one without being able to formally call them… we would just, if an ambulance came in, we were like, ‘please, thank you for unloading this patient and here’s another one, please go to the other hospital.’ It was really bad for them and us as well. It was very hectic.”

#### Reduced Coordination with Ambulance Services

Another significant problem identified by respondents was the challenge of communicating with ambulance services. Respondents noted that even when they identified a patient that needed to be transferred, they were often unable to secure an ambulance to transfer the patient for various reasons. As one EM physician noted, “We couldn’t communicate with the ambulance systems… you couldn’t coordinate for a patient to be transferred through our ambulance. At the same time, how are you going to discharge somebody who has an amputation and needs treatment.” Another problem was ambulance services refusing to transfer patients unless they first received cash payment. One EM physician noted, “A big thing would be our partnership with the ambulances. For it to be stronger and for us to have an ambulance with us, at least right there, 24 hours… [then] we don’t have to wait until paramedics, because they’re from private companies… it’s very odd and they basically want $250 up front instead of transferring the patient that’s going to die in their face.” The problem was that the hospital could not communicate to the ambulance services that the patient’s insurance would cover ambulance, so private companies demanded cash payment. Another respondent noted, “Once we were able to move patients, we didn’t have the transportation, the ambulances… while we worked to maintain the health and conditions of the patients, the ambulance companies didn’t want to accept any medical plans.”

#### Poor Response Coordination with Government

Many respondents noted poor and inefficient coordination with the Puerto Rican government. As one administrator noted, “When everything happened, communication with agencies was non-existing. We had to go to COE (Centro de Operacion de Emergencias, Emergency Operation Center), which was in San Juan. There, the government integrated all agencies into a single place, and that’s how we all communicated. We had to go there to ask for help and determine what we were going to do.” In the case of Hurricane Maria, vertical communication was absent or significantly slowed.

#### Community Trust in Hospital and Healthcare System

Respondents identified two themes that they felt contributed to more resilient response following Hurricane Maria. The first theme identified as contributing to resilience was the large degree of community trust in this PR Community Hospital in the weeks following Hurricane Maria. As one FM physician mentioned, “This hospital is known in the community as the ‘Hospital of the area,’ so the communities surrounding us know this is the hospital they go to when something happens. This is not just an emergency hospital, but a place the community comes for everything because they know this place provides all the services. The hospital published a group of messages in the newspapers to communicate that it was open and able to provide services. That’s another way for the community to know they could come and would be assisted.” The other theme was the relationship healthcare workers had cultivated with the community. This relationship aided resilience after the Hurricane and made patients confident they could rely on Puerto Rican health providers.

### Adaptive

#### Organized Staffing Adjustments

Respondents noted that the UPR Community Hospital staff was remarkably flexible in the days and weeks after the Hurricane. The UPR Community Hospital increased staff and work hours in the weeks following the Hurricane and the physicians were able to adapt without problem. As one EM physician states, “They did a really good job right after making sure that we had enough staff. We worked doubles. Not double shifts, but we had a double coverage. The nursing staff was really amazing about that. We have people that live really far away. We had one nurse who actually lost her whole house and they were really supportive about getting her new uniforms and helping her get to work. We rallied together. They were really helpful in that sense.” Despite the difficulties in coordinating care and infrastructure, the Puerto Rican health care staff was able to effectively adapt to the circumstances in a manner that cultivated resilience.

## Discussion

It has been over two years since Hurricane Maria made landfall in Puerto Rico, and the impact of the Hurricane can still be felt today. Various studies have shown the widespread devastation that Hurricane Maria had on the people of Puerto Rico: taking nearly 5,000 lives, devastating the health system, and destroying the physical infrastructure of the island [2]. Our study focuses not on the magnitude of the destruction, but on the specific factors intrinsic to the Puerto Rican health system that may have predisposed the island’s population to experiencing more severe health outcomes. Specifically, we performed a ‘critical case’ case study guided by health system resilience framework using semi-structured interviews with health care providers at a UPR Community Hospital. **Figure 1** uses the Kruk framework of health system resilience to explain resilience in Puerto Rico following Hurricane Maria.

**Figure 1:**
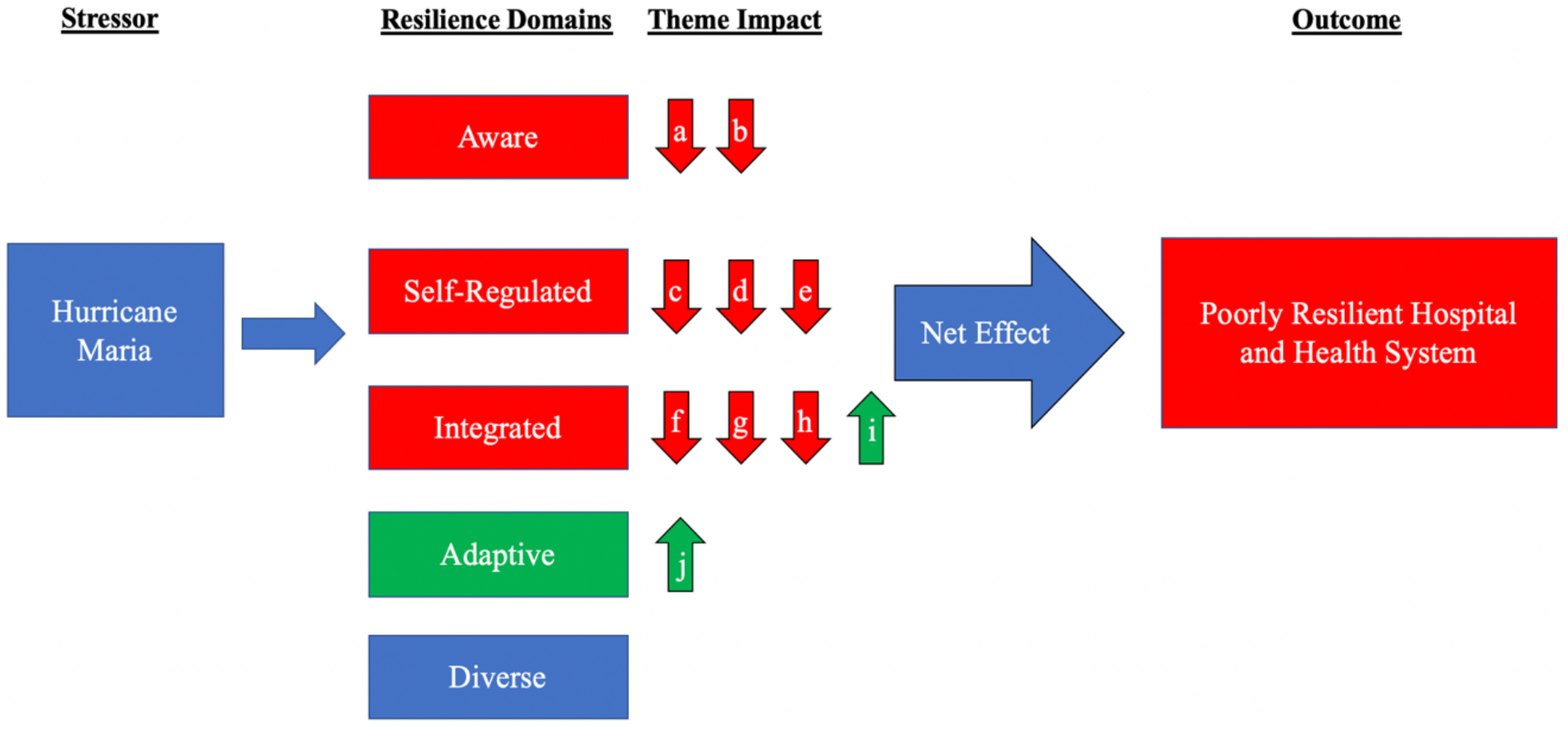
Model of Aggregate Individual Domain Effect on Health System Resilience. ^a^Recognizing Capacity of Back-Up Generators Before Hurricane Maria, Understanding Necessity to Transfer ICU patients Before Hurricane Maria, ^c^Treating Patients with End-Stage Renal Disease Patients, ^d^Access to Medications for Patients with Chronic Conditions, ^e^Admitting New Patients, ^f^Coordinating Care with Tertiary Care Centers, ^g^Coordinating with Ambulance Services, ^h^Response Coordination with Government ^i^Community Trust in Hospital and Healthcare System, ^j^Staffing Adjustment

This study identified key factors within the Aware, Self-Regulated, and Integrated domains that were especially weak and were the key factors that led to a poorly resilient hospital and health system. The findings pointed to specific areas that can be targeted to improve the resilience of the Puerto Rican health system and can be adapted to other health systems. First, ensuring that key hospitals have adequate back-up generators is imperative. The lack of back-up generators resulted in an interruption of health services that spilled over into other domains of resilience. Puerto Rico and other nations with limited resources constantly facing threats of natural disasters must prioritize funding back-up generators at key hospitals.

Second, our study corroborated the findings of previous studies that the patients most at risk for adverse health outcomes immediately after a natural disaster are patients with chronic conditions. These patients rely on constant access to medications and health services for their livelihood. The most dramatic example of these patients is those with ESRD, but also included in this patient population are those with heart failure, chronic psychiatric conditions, and COPD patients. Following Hurricane Maria, the health system was overwhelmed by these patients presenting acutely decompensated conditions requiring emergency management. To ensure that these patients are adequately cared for, pharmacies and dialysis centers also need to be prioritized as healthcare facilities with adequate back-up generators and resources.

Third, there must be improved measures for communication both horizontally and vertically. Horizontally, a system must be put in place to transfer critically ill patients to appropriate facilities with back-up generators. Vertically, the government must have a system in place that ensures emergency medical services are guaranteed reimbursement during a natural disaster.

Our study did have limitations. Most significantly, this is a qualitative case study with a limited number of participants. Because this was a single-institution case study our findings may not be generalizable. However, the study was designed to sample a ‘critical case’ scenario, and while our findings cannot be generalizable to all healthcare systems, a *logical generalization* can be made that, similar to the UPR Community Hospital, other PR hospitals *logically* may have had similar or worse experiences. In addition, although we interviewed a range of providers and administrators, we did reach data saturation and our respondents were able to speak not just to about the UPR Community Hospital but also to its interplay within Puerto Rico and the healthcare system as a whole. That being said, future resilience framework studies should incorporate the perspectives to health providers and other institutions and to public health professionals.

In September 2017, the people of Puerto Rico were devastated by Hurricane Maria. When there is a stress to a health system, the population will inevitably experience some heightened degree of social disruption, morbidity and mortality. As we have seen with the outbreak of COVID-19, it is as important as ever that both developed and developing countries are fortified with resilient health systems so that when an inevitable stress occurs, social disruption and excess morbidity can be minimized. Unfortunately, the degree of devastation experienced by the people of Puerto Rico after Hurricane Maria demonstrates a poorly resilient health system [10; 18]. Yet, up to this point, there has not been a study that examines the specific aspects of the Puerto Rican health system that resulted in this poorly resilient response. Our critical case study is the first to identify aspects of the Puerto Rican health system that led to a poorly resilient response and can be targeted by the Puerto Rican Public Health officials to improve the resilience before the next disaster hits. With limited government funding and global warming resulting in more devastating storms each year, it is imperative that Puerto Rico and other resource limited nations focus their investments appropriately in order to cultivate health system resilience and protect their populations against the next inevitable health system shock.

## Data Availability

N/A

## Acknowledgements

We would like to thank the staff of the UPR Community hospital for taking the time to participate in this study. ***We plan to report the results of this study to all participants***.

## Competing Interests and Funding

All authors have completed the ICMJE uniform disclosure form and declare no financial support or relationships with any organizations that might have an interest in the submitted work in the previous three years, no other relationships or activities that could appear to have influenced the submitted work. This work was supported by an NIH R01 HL134776 and R01 HL139664 to V. de Jesus Perez. C. Rios was supported by a Stanford Medical Scholars Program. The funding sources were not involved in the design, data capture, analysis of drafting of the manuscript.

## Contributors and Guarantor Information

This project was carried out by C. Rios as the topic of his Stanford Medical Scholars research project under the supervision of Dr. de Jesus Perez. Details on the scope of the Stanford Medical Scholars Program can be found at https://med.stanford.edu/medscholars.html. C. Rios was primarily responsible for designing the study, implementing the resilience framework, conducting the interviews, analyzing the data and drafting the manuscript. E. Ling provided input on adapting the Kruk resilience framework for this study. Drs. R. Rivera-Gutierrez and J. Gonzalez assisted in translating the framework and identifying participants at the UPR Community Hospital. J. Bruce and Dr. M. Barry were involved in data analysis and manuscript preparation. Dr. de Jesus Perez was involved in the conception, design, organization of resources, data analysis and manuscript preparation. As the corresponding author, Dr. de Jesus Perez attests that all listed authors meet authorship criteria and that no others meeting the criteria have been omitted. Dr. de Jesus Perez confirms that the manuscript is an honest, accurate and transparent account of the study being reported and that no important aspects of the study have been omitted.

## Notes

### Competing Interest Statement

The authors have declared no competing interest.

### Author Declarations

The Stanford Institutional Review Board approved all research procedures as an exempt study

## References

[1] Santos-Burgoa, Ascertainment of the Estimated Excess Mortality From Hurricane María in Puerto Rico, The George Washington University Milken Institute School of Public Health, 2018.

[2] N. Kishore, D. Marques, A. Mahmud, M.V. Kiang, I. Rodriguez, A. Fuller, P. Ebner, C. Sorensen, F. Racy, J. Lemery, L. Maas, J. Leaning, R.A. Irizarry, S. Balsari, and C.O. Buckee, Mortality in Puerto Rico after Hurricane Maria. N Engl J Med 379 (2018) 162–170.

[3] M.E. Kruk, E.J. Ling, A. Bitton, M. Cammett, K. Cavanaugh, M. Chopra, F. El-Jardali, R.J. Macauley, M.K. Muraguri, S. Konuma, R. Marten, F. Martineau, M. Myers, K. Rasanathan, E. Ruelas, A. Soucat, A. Sugihantono, and H. Warnken, Building resilient health systems: a proposal for a resilience index. BMJ 357 (2017) j2323.

[4] M.E. Kruk, M. Myers, S.T. Varpilah, and B.T. Dahn, What is a resilient health system? Lessons from Ebola. Lancet 385 (2015) 1910–2.

[5] E.J. Ling, E. Larson, R.J. Macauley, Y. Kodl, B. VanDeBogert, S. Baawo, and M.E. Kruk, Beyond the crisis: did the Ebola epidemic improve resilience of Liberia’s health system? Health Policy Plan 32 (2017) iii40–iii47.

[6] S. Abimbola, and S.M. Topp, Adaptation with robustness: the case for clarity on the use of ‘resilience’ in health systems and global health. BMJ Glob Health 3 (2018) e000758.

[7] K. Blanchet, S.L. Nam, B. Ramalingam, and F. Pozo-Martin, Governance and Capacity to Manage Resilience of Health Systems: Towards a New Conceptual Framework. Int J Health Policy Manag 6 (2017) 431–435.

[8] J. Kutzin, and S.P. Sparkes, Health systems strengthening, universal health coverage, health security and resilience. Bull World Health Organ 94 (2016) 2.

[9] J. Greenstein, J. Chacko, B. Ardolic, and N. Berwald, Impact of Hurricane Sandy on the Staten Island University Hospital Emergency Department. Prehosp Disaster Med 31 (2016) 335–9.

[10] L.C. Pullen, Puerto Rico after Hurricane Maria. Am J Transplant 18 (2018) 283–284.

[11] J. Mattei, M. Tamez, C.F. Rios-Bedoya, R.S. Xiao, K.L. Tucker, and J.F. Rodriguez-Orengo, Health conditions and lifestyle risk factors of adults living in Puerto Rico: a cross-sectional study. BMC Public Health 18 (2018) 491.

[12] X. Li, J. Huang, and H. Zhang, An analysis of hospital preparedness capacity for public health emergency in four regions of China: Beijing, Shandong, Guangxi, and Hainan. BMC Public Health 8 (2008) 319.

[13] R.K. Lin, Applications of Case Study Research, SAGE Publications, the Univeristy of California, 1993.

[14] M.Q. Patton, Qualitative Research & Evaluation Methods, SAGE, 2002.

[15] D.V. 8.0.35, web application for managing, analyzing, and presenting qualitative and mixed method research, SocioCultural Research Consultants Los Angeles, CA, 2018.

[16] B.C. O’Brien, I.B. Harris, T.J. Beckman, D.A. Reed, and D.A. Cook, Standards for reporting qualitative research: a synthesis of recommendations. Acad Med 89 (2014) 1245–51.

[17] A. Strauss, Corbin, J. M., Basic of Qualitative Research: Techniques and Procedures for Developing Grounded Theory, SAGE Publications, 1998.

[18] J. Roman, Hurricane Maria: A Preventable Humanitarian and Health Care Crisis Unveiling the Puerto Rican Dilemma. Ann Am Thorac Soc 15 (2018) 293–295.

[19] A.H. Mokdad, G.A. Mensah, S.F. Posner, E. Reed, E.J. Simoes, M.M. Engelgau, D. Chronic, and G. Vulnerable Populations in Natural Disasters Working, When chronic conditions become acute: prevention and control of chronic diseases and adverse health outcomes during natural disasters. Prev Chronic Dis 2 Spec no (2005) A04.

[20] R.M. Hakim, and J.M. Lazarus, Initiation of dialysis. J Am Soc Nephrol 6 (1995) 1319–28.

